# Feasibility characteristics of wrist-worn fitness trackers in health status monitoring for post-COVID patients in remote and rural areas

**DOI:** 10.1101/2023.12.05.23299499

**Authors:** Madeleine Wiebe, Marnie Mackay, Ragur Krishnan, Julie Tian, Jake Larsson, Setayesh Modanloo, Christiane Job Mcintosh, Melissa Sztym, Gail Elton-Smith, Alyssa Rose, Chester Ho, Andrew Greenshaw, Bo Cao, Andrew Chan, Jake Hayward

**Affiliations:** Faculty of Medicine and Dentistry, University of Alberta; Department of Psychiatry, Faculty of Medicine and Dentistry, University of Alberta; Department of Electrical and Computer Engineering, Faculty of Engineering University of Alberta; Neurosciences, Rehabilitation and Vision Strategic Clinical Network, Alberta Health Services; Covenant Health Rural Health Services; Glenrose Rehabilitation Hospital; Department of Physical Medicine and Rehabilitation, Faculty of Medicine and Dentistry, University of Alberta; Department of Emergency Medicine, Faculty of Medicine and Dentistry, University of Alberta

## Abstract

**Introduction:** Common, consumer-grade biosensors mounted on fitness trackers and smartwatches can measure an array of biometrics that have potential utility in post-discharge medical monitoring, especially in remote/rural communities. The feasibility characteristics for wrist-worn biosensors are poorly described for post-COVID conditions and rural populations.

**Methods:** We prospectively recruited patients in rural communities who were enrolled in an at-home rehabilitation program for post-COVID conditions. They were asked to wear a FitBit Charge 2 device and biosensor parameters were analyzed (e.g. heart rate, sleep, and activity). Electronic patient reported outcome measures (E-PROMS) for mental (bi-weekly) and physical (daily) symptoms were collected using SMS text or email (per patient preference). Exit surveys and interviews evaluated the patient experience.

**Results:** Ten patients were observed for an average of 58 days and half (N=5) were monitored for 8 weeks or more. Five patients (50%) had been hospitalized with COVID (mean stay = 41 days) and 4 (36%) had required mechanical ventilation. As baseline, patients had moderate to severe levels of anxiety, depression, and stress; fatigue and shortness of breath were the most prevalent physical symptoms. Four patients (40%) already owned a smartwatch. In total, 575 patient days of patient monitoring occurred across 10 patients. Biosensor data was usable for 91.3% of study hours and surveys were completed 82.1% and 78.7% of the time for physical and mental symptoms, respectively. Positive correlations were observed between stress and resting heart rate (r=0.360, p<0.01), stress and daily steps (r=0.335, p<0.01), and anxiety and daily steps (r=0.289, p<0.01). There was a trend toward negative correlation between sleep time and physical symptom burden (r=-0.211, p=0.05). Patients reported an overall positive experience and identified the potential for wearable devices to improve medical safety and access to care. Concerns around data privacy/security were infrequent.

**Conclusions:** We report excellent feasibility characteristics for wrist-worn biosensors and e-PROMS as a possible substrate for multi-modal disease tracking in post-COVID conditions. Adapting consumer-grade wearables for medical use and scalable remote patient monitoring holds great potential.

## Introduction

Recovery from COVID can be long and unpredictable. Some patients return to premorbid health quickly while others endure a debilitating and poorly understood recovery path lasting months to years, termed ‘long-COVID’, comprising both mental and physical symptoms (reviewed in Davis et al., 2023 and Koc et al., 2022). Traditional, clinic-based methods of disease assessment are episodic, limiting their utility in a complex and undulating disease course such as post-COVID. New approaches are needed that can track longitudinal physiologic changes at the individual patient level.

Wearable devices may be ideal tools for studying complex diseases and post-COVID conditions (reviewed in Smuck et al., 2021). Biosensor metrics have been shown able to predict health outcomes for a range of chronic diseases including congestive heart failure, COPD, hypertension, diabetes, and more (Channa et al., 2021; Khondakar & Kaushik, 2022; Rodriguez-León et al., 2021; Singhal & Cowie, 2020). More recently, smartwatches have been used to detect pre-symptomatic COVID infections (Mishra et al., 2020). Common and affordable smartwatches and fitness trackers might support remote patient monitoring (RPM) if the emergent data is high-enough quality to inform safe decision making (Iqbal et al., 2021, Kwok et al., 2021; Lu et al., 2020, Kang & Exworthy, 2022). Remote patient monitoring is particularly important for patients living in remote and rural areas where in-person care can be limited (Fraser et al., 2022; Liao et al., 2019; Canali et al., 2022).

Feasibility data for wearable devices varies with population, technology, and monitoring protocol and little has been published for post-COVID conditions, especially in rural settings. Our study describes the feasibility of using common, consumer-grade wrist-worn biosensors (e.g., FitBit fitness trackers) for disease parameter tracking in post-COVID conditions in a rural community, including both technical aspects (e.g. data quality/completeness) and the patient experience.

## Materials and Methods

### Patient Cohort

Patients were recruited from an early supported discharge (ESD) program in Camrose, Alberta, Canada, which has a population of just under 21,000 residents. The ESD program was originally designed to support patients recovering from acute stroke (Chouliara et al., 2023); however, during the COVID-19 pandemic, it was adapted for COVID-19. Patients were referred to the program either directly from hospital (at discharge) or through community clinics. The ESD team is a rehabilitation team that delivers intensive rehabilitation programs in the home using both in-person visits and telemedicine (video or phone). The team typically includes occupational therapy, physical therapy, speech-language pathology, social work, nursing, therapy assistants, recreation therapy and a psychologist. Patients were also recruited through a virtual post-covid pilot program and were located throughout Alberta, primarily in rural settings. Patients were referred to this program through the Rehabilitation Advice Line. Rehabilitation in this program was provided solely via telemedicine.

All ESD patients were screened for eligibility at program start. Eligible patients were ambulatory, with mild to moderate functional impairment and sufficient cognitive capacity to participate in the program. Excluded patients were those without internet connectivity, who were unable to complete surveys (ex. language and speech deficits, non-English-speaking), or for whom device retrieval was deemed to be unlikely by the study investigators.

### Data Sources

Data sources included: 1) wearable devices, 2) health records, 3) digital surveys, and 4) patient interviews. Health record data comprised diagnostic, medication, and procedure codes occurring in the year before enrollment and during the observation period.

### Devices

FitBit Charge 2 devices were provided to each patient at enrollment. The FitBit Charge 2 is a wrist-worn fitness tracker that includes a triaxial accelerometer, an altimeter, and an optical heart rate tracker. Parameters collected included activity (steps per day), heart rate (beats per minute), and duration (minutes) and quality (% deep) of sleep. Data was sent via Bluetooth to a patient-owned smartphone, uploaded to the FitBit cloud database and then extracted through the third-party platform (Fitabase; https://www.fitabase.com/).

### Monitoring Protocol

Patients received an orientation to the device on study enrollment and were instructed to wear it as much as possible, up to 24 hrs a day (except when charging). There were no specific instructions on how to use device data and patients were free to share data with the clinical team if desired; however, data was not directly transmitted to the clinical team. If the patient had questions about the device or technical issues, a research nurse was available for support by phone during daytime hours.

### Surveys

Patients received digital surveys via SMS text or email (per patient preference) using the RedCap (Harris et al., 2008) survey platform. Baseline health status surveys included measures of mental and physical health and a technology experience survey (Comprising two sections: Tech comfort (7-items, avg score [0-5]), Health Literacy (4-items, avg score [0-5]) adapted from the Telehealth Usability Questionnaire (TUQ) (Parmanto et al., 2016). During the follow-up period physical and mental symptom surveys were delivered daily and bi-weekly, respectively. English versions of validated self-reported screening scales were used to measure severity of stress, anxiety, and depression symptoms, including the Perceived Stress Scale (PSS), a 10-item questionnaire with a Cronbach’s alpha of >0.70 used to assess level of stress in the previous month (PSS; PSS score ≥14 indicates moderate or high stress) (Cohen et al., 1983); the Generalized Anxiety Disorder 7-item (GAD-7) scale, a 7-item questionnaire with a Cronbach’s alpha of 0.92 and used to assess the self-reported levels of anxiety in respondents in the two weeks prior to assessment (GAD-7 score ≥10 indicates likely generalized anxiety disorder [GAD]) (Spitzer et al., 2006); and the Patient Health Questionnaire-9 (PHQ-9) a 9-item questionnaire with a Cronbach’s alpha of 0.89 and used to assess the severity of depression symptoms (for PHQ-9; a score ≥10 indicates likely major depressive disorder (MDD) (Kroenke et al., 2001).

Detailed description of physical symptom surveys and results can be found in the supplementary materials, Table S1. In short, 18 total symptom scales (6-point scale, ‘absent’ [1] to ‘very severe’ [6]) were grouped by body system (constitutional, gastrointestinal, neurologic, respiratory, cardiovascular). At discharge, patients received an exit survey that included feedback on the devices, also adapted from the TUQ.

### Interviews

At the conclusion of the study patients were approached for interview. Questions focused on technology usability, acceptance, and perceived barriers to use, such as data privacy/security. Author MW completed interviews virtually using Zoom and curated auto-generated transcriptions.

## Study Outcomes

### Primary Outcome: Protocol Adherence

Our primary outcome was protocol adherence, defined as device wear time (% of minutes with analyzable heart rate data) and survey completion rates. Patient interviews and survey responses added context for this outcome, exploring patient perceptions and experience.

### Secondary Outcomes: Biosensor Parameter Correlations

Secondary outcomes were associations between 1) the primary outcome (adherence) and patient characteristics (e.g., age, sex, symptom severity, technology readiness measures, and time under observation), and 2) device biometrics (activity/HR/sleep) and disease outcomes (physical and mental symptoms).

### Statistical Analysis

Descriptive statistics (mean and standard deviation [SD]) were calculated for primary and secondary outcomes. We used Pearson correlation coefficients and weekly aggregated data to explore secondary outcomes. We performed a within-subject longitudinal analysis and paired t-tests to evaluate for changes in symptom severity over time, comparing weeks 1-4 and weeks 8+ for those subjects observed for more than 8 weeks (N=5).

### Qualitative Analysis

We conducted thematic analysis of interview transcripts using a combined deductive and inductive approach, aided with NVIVO software. Two team members (MW and JL) reviewed original transcripts to identify themes of interest and compared results. Where there was disagreement, consensus was obtained through discussion. Smaller themes were sequentially grouped into larger categories until core themes were identified.

### Funding and Ethics

This study was registered and approved by the University of Alberta Research Ethics Board (Pro00113943).

## Results

### Participant characteristics

Between November 2021 and May 2022, 18 patients were approached for enrollment; three declined and one patient was ineligible due to lack of wireless connectivity. After enrollment, 3 of 14 patients withdrew consent, and 1 was removed due to technical issues with their device, leaving 10 patients for the final analysis. Seven patients consented to an interview. The overall recruitment rate was therefore 61.1%.

Table 1 shows baseline patient characteristics. Mean age was 53 years (SD 15.0) and 80% (8/10) of patients were female. Five (50%) had been hospitalized with COVID (average length of stay = 41 days) and four (40%) had required ventilatory support in the ICU. The remaining 50% were referred from community clinics. On average, patients had fewer than one documented medical condition prior to their COVID-19 diagnosis and half (5/10) owned their own smartwatch. The most common physical symptoms were ‘constitutional’ (ex. fatigue, poor sleep), followed by ‘respiratory’ and ‘cardiovascular’. Average mental health symptoms scores were GAD-7 = 15.7 (severe anxiety), PHQ-9 = 14.8 (moderate depression) and PSS = 22.7 (moderate stress). Patients had moderate to high levels of familiarity with technology (ex. comfort with technology = 3.6/5; health literacy = 3.7/5). Detailed patient characteristics, including comorbidities and medications, are shown in Supplementary Table S1 and Figures S1 and S2.

**Table 1.** Baseline patient characteristics (N=10)

| **Demographics** |  |
| --- | --- |
| Age (mean [SD]) | 53 [15] |
| Female Sex no. (%) | 8 (80%) |
| Hospitalized no. (%) | 5 (50%) |
| Days in hospital (mean [SD])* | 41.0 [21.1] |
| ICU stay no. (%) | 4 (40%) |
| Ventilator no. (%) | 4 (40%) |
| Number of days in ESD (mean [SD]) | 58 [117] |
| Home oxygen (%) | 1 (10%) |
| Comorbidities per patient (mean [SD]) | 0.8 [0.8] |
| Medications per patient (mean [SD]) | 4.4 [3.1] |
| **Familiarity with Technology** |  |
| Already own a smartwatch (%) | 4 (40%) |
| Technology comfort (7-items, mean score [0-5]) | 3.6 [1.4] |
| Health Literacy (4-items, mean score [0-5]) | 3.7 [1.1] |
| **Physical symptoms (mean score [0-4])** |  |
| Constitutional (6 symptoms) | 1.43 [0.83] |
| Gastrointestinal (3 symptoms) | 0.31 [0.55] |
| Neurological (3 symptoms) | 0.61 [0.62] |
| Respiratory (4 symptoms) | 0.90 [0.55] |
| Cardiovascular (2 symptoms) | 0.68 [0.90] |
| **Mental health symptoms (mean score [SD])** |  |
| General Anxiety Disorder-7 (out of 21) | 15.7 [1.3] |
| Patient Health Questionnaire-9 (out of 27) | 14.8 [1.3] |
| Perceived Stress Scale (out of 40) | 22.7 [1.4] |

\* hospitalized patients (N=5)

### Descriptive analysis

Table 2 summarizes descriptive statistics for primary adherence outcomes (group aggregate). Overall, 575 patient days of patient monitoring occurred across 10 patients, patients were observed for 58 days on average and half (N=5) were monitored for 8 weeks or more. For the primary outcome (adherence), heart rate data was available for 91.3% of study hours and physical and mental symptom surveys were completed 82.1% and 78.7% of the time, respectively. Overall physical symptom burden was generally mild with an average overall rating of 0.8/4; constitutional and respiratory symptoms being the most frequent. Average group activity, sleep duration and quality (proportion time in deep sleep) were highest in weeks 4-8. Average resting heart rate was relatively constant over time. To test for within-subject effects, we compared weeks 1-4 and weeks 8+ for those patients staying longer than 8 weeks (Table S2). Except for respiratory symptoms (mean difference –0.23, p=0.03), temporal changes did not reach statistical significance.

**Table 2.** Reported health parameters and survey outcomes across the clinical course.

| Category | Weeks 1-4 (N= 10)<br>Average [SD] | Weeks 4-8 (N= 9)<br>Average [SD] | Weeks 8+ (N= 5)<br>Average [SD] |
| --- | --- | --- | --- |
| % Wear time<br>(minutes with<br>heart rate data) | 91.5 [5.4] | 92.9 [4.4] | 88.1 [5.7] |
| Sleep (hours) | 5.7 [2.3] | 7.0 [2.0] | 5.6 [3.2] |
| Deep sleep (%) | 13.3 [10.0] | 17.4 [7.5] | 15.9 [8.2] |
| Resting heart rate | 68.7 [7.6] | 66.1 [9.5] | 66.7 [8.9] |
| % Completion for surveys |  |  |  |
| Physical | 82.8 [12.3] | 81.1 [18.1] | 82.5 [11.2] |
| Mental | 92.1 [15.3] | 81.0 [23.1] | 47.9 [36.1] |
| Physical symptoms (mean [SD]; range 0-4) |  |  |  |
| All | 0.8 [0.6] | 0.7 [0.5] | 0.7 [0.4] |
| Constitutional | 1.0 [0.8] | 0.9 [0.8] | 1.1 [0.7] |
| Neuro | 0.6 [0.6] | 0.6 [0.6] | 0.8 [0.8] |
| Gastrointestinal | 0.3 [0.6] | 0.2 [0.5] | 0.3 [0.5] |
| **Respiratory** | 0.9 [0.6] | 0.8 [0.6] | 0.8 [0.5] |
| **Cardiovascular** | 0.7 [0.9] | 0.6 [0.9] | 0.3 [0.6] |
| **Mental health symptoms** |  |  |  |
| **Mood (out of 27)** | 20.0 [8.0] | 19.3 [9.0] | 16.4 [7.8] |
| **Anxiety (out of 21)** | 16.2 [6.8] | 15.8 [6.8] | 12.4 [6.4] |
| **Stress (out of 40)** | 30.5 [6.1] | 28.5 [4.9] | 26.6 [5.5] |
| **Activity: Steps** |  |  |  |
| **Daily count** | 3042 [1606] | 4330 [3338] | 3284 [2249] |
| **Activity: Intensity (minutes/day)** |  |  |  |
| **Low** | 127.3 [30.2] | 156.6 [71.7] | 121.6 [74.3] |
| **Moderate** | 6.4 [6.6] | 12.6 [16.9] | 12.4 [20.3] |
| **High** | 4.8 [6.2] | 5.2 [6.6] | 5.6 [6.1] |

### Group-level associations

Survey response rates were correlated with device wear time (r=0.67, p=0.03), however there were no statistically significant associations between wear-time and other patient characteristics (e.g. demographics, symptoms, previous technology experience or time under observation; Table S3). Table 3 shows group-level associations between biosensor metrics (weekly aggregate). Resting heart rate was positively correlated with stress (r=0.360, p<0.01); step count was correlated with anxiety (r=0.289, p<0.01) and stress (r=0.335, p<0.01). Correlations for sleep time were not statistically significant, however trends suggested negative correlations with physical symptoms (r=-0.211, p=0.05)) and anxiety (r=-0.186, p=0.09)) and a positive correlation with stress (r=0.208, p0.06)).

**Table 3.** Correlation coefficients between FitBit data outputs and patient symptoms.

| Survey parameter | Resting Heart Rate | Steps | Sleep Time |
| --- | --- | --- | --- |
| Physical Symptoms | 0.011 (P=0.920) | 0.116 (P=0.30) | -0.211 (P=0.05) |
| Anxiety (GAD-7) | 0.205 (P=0.070) | 0.289 (P<0.01) | -0.186 (P=0.09) |
| Mood (PHQ-9) | 0.148 (P=0.189) | 0.183 (P=0.10) | 0.004 (P=0.97) |
| Stress (PSS) | 0.360 (P<0.01) | 0.335 (P<0.01) | 0.208 (P=0.06) |

### Individual sensor data: case examples

Figure 2 shows data for two patients, illustrating the complexity and heterogeneity of individual data. At enrollment, Patient A experienced marked physical symptoms, a high resting heart rate (79-83 bpm) and low daily step counts (approx. 1km). Over time, their physical symptoms improved, resting heart rate decreased, and daily activity increased. In contrast, Patient B’s physical symptoms remained prominent throughout observation. Their resting heart rate was lower than Patient A (46-54 bpm) and didn’t significantly change over time. Their activity levels didn’t show a clear temporal change. Mental health symptoms were prominent in both patients.

**Figure 1.**
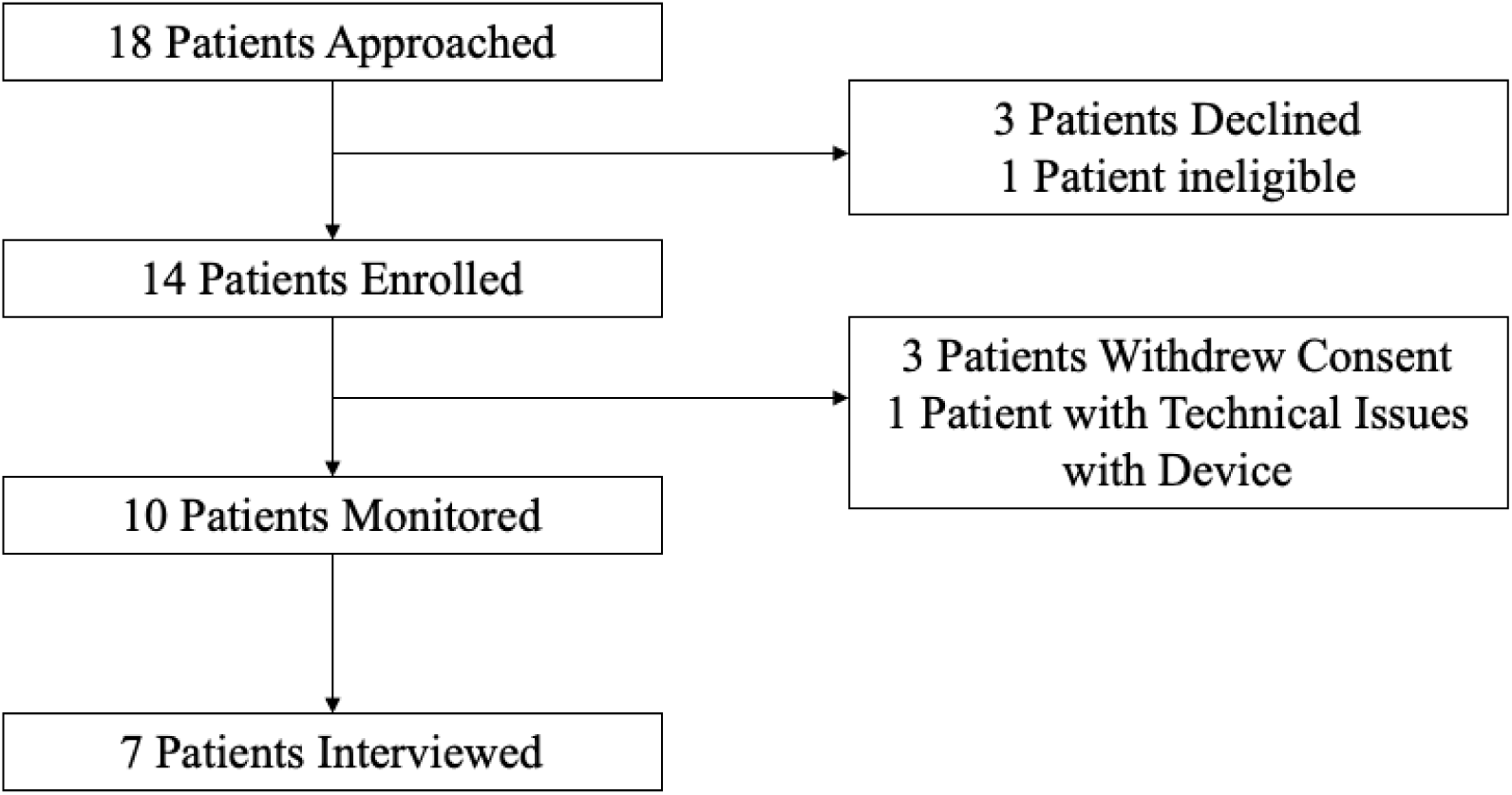
Flow diagram of study recruitment.

**Figure 2.**
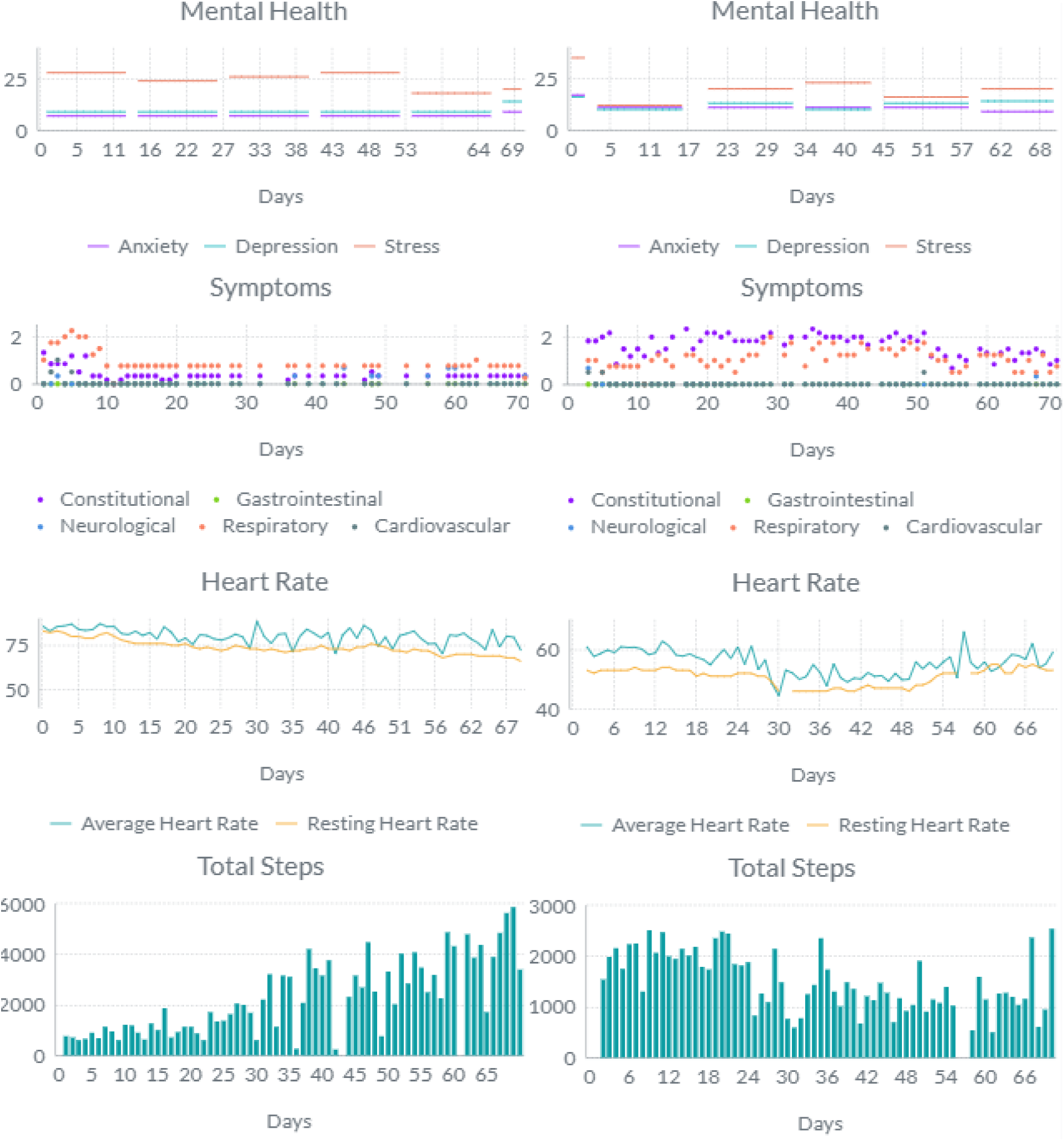
Sample longitudinal data for Patient A (left) and B (right), including results from symptom surveys, heart rate and total daily number of steps.

### Surveys results

Figures 2 and 3 display survey results; additional survey data is presented in supplementary materials (Figure S1). Proportions herein represent overall levels of agreement (agree or strongly agree) vs. disagreement (disagree or strongly disagree). Of those who responded, the majority liked using the FitBit system (5/8 [63%]), found it easy to learn (6/8 [86%]) and simple to use (7/7 [100%]); no patients found the device interfered with their lifestyle and only one (13%) found the device uncomfortable. Three-quarters (6/8 [75%]) of the patients found that wearing a device made them feel safer and 4/8 (50%) felt it helped them to better understand their disease. A minority (2/8 [25%]) used the device to help decide when to seek medical care and no patients (0/8) reported that the devices caused anxiety.

**Figure 3.**
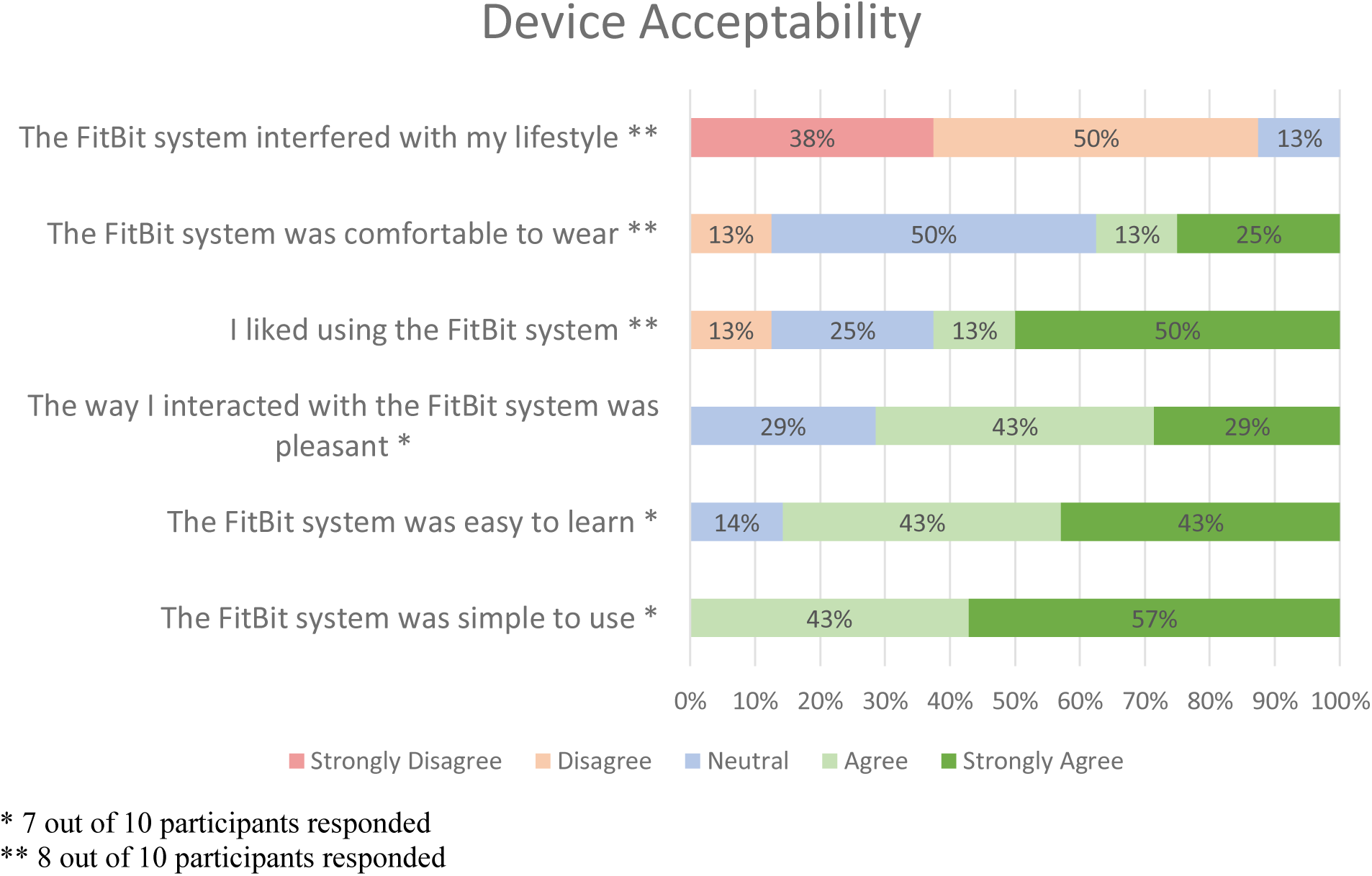
Survey results for questions pertaining to device acceptability with answers ranging from strongly disagree to strongly agree.

**Figure 4.**
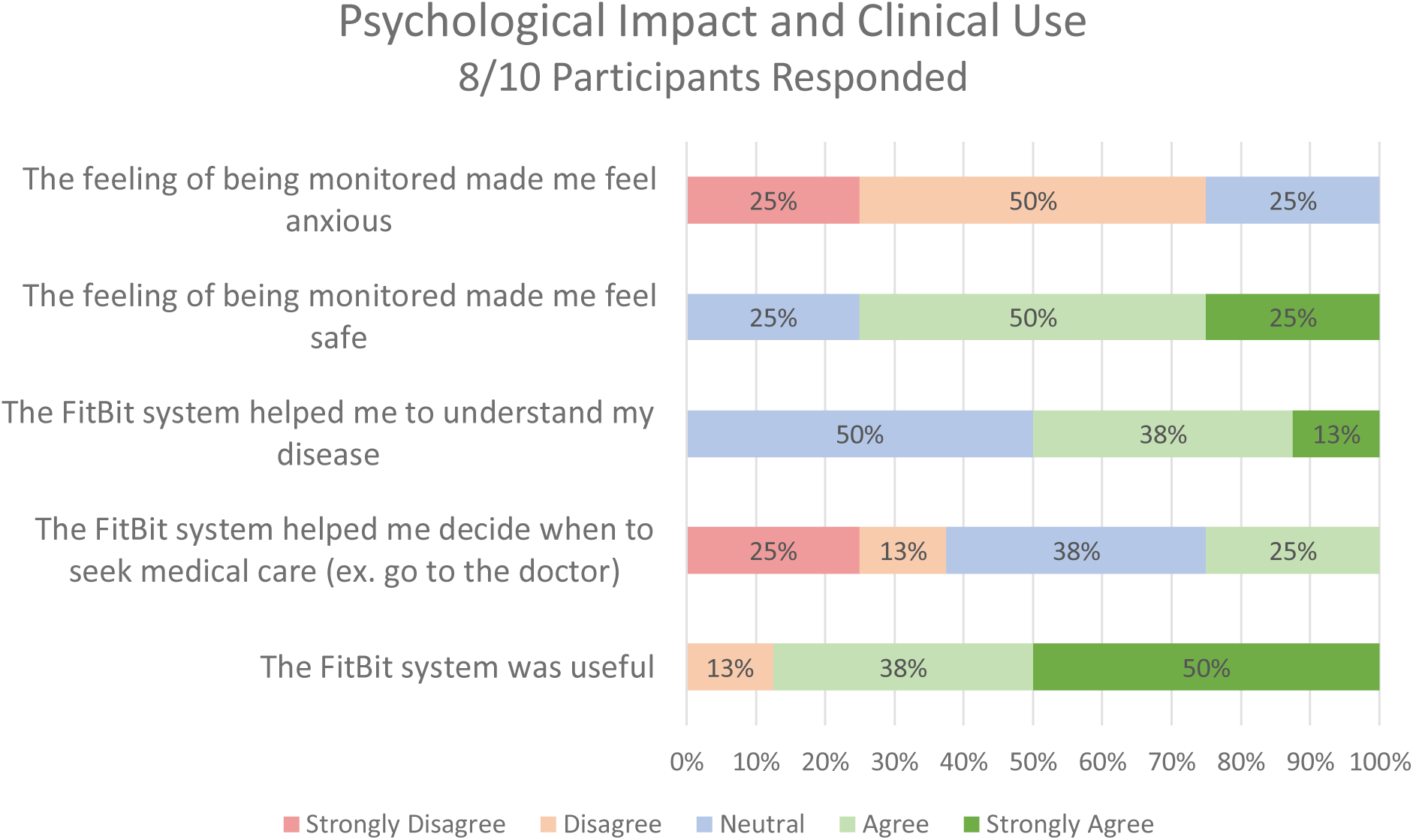
Survey Results for questions pertaining to psychological impact and clinical use with answers ranging from strongly disagree to strongly agree.

### Theme analysis of interview transcripts

Two male and five female patients completed exit interviews. Key themes are summarized below, and representative quotes are presented in Table 4.

**Table 4.**
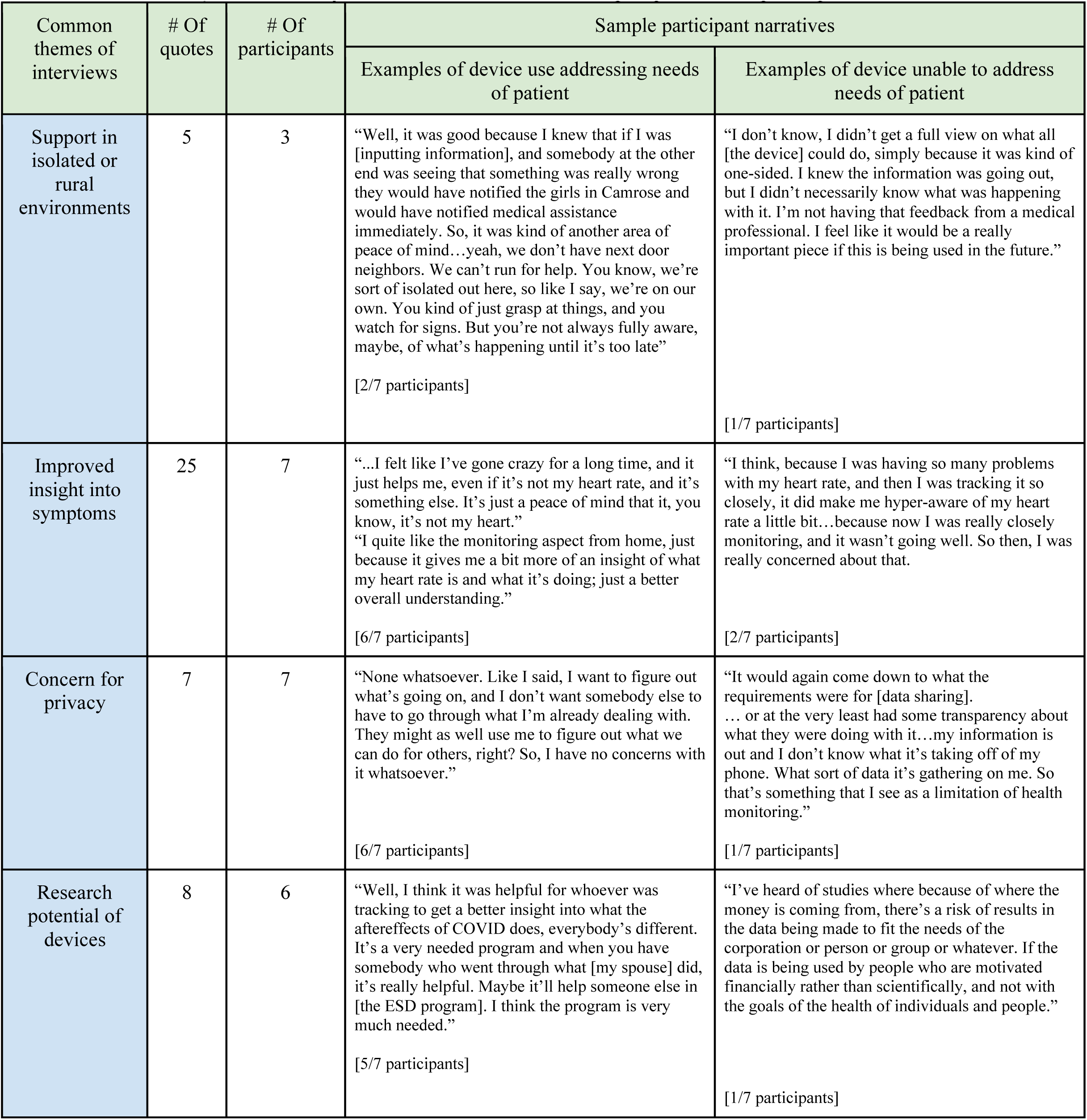
Qualitative analysis of exit interviews with sample quotes from participants.

| Common themes of interviews | # Of quotes | # Of participants | Sample participant narratives |  |
| --- | --- | --- | --- | --- |
|  |  |  | Examples of device use addressing needs of patient | Examples of device unable to address needs of patient |
| Support in isolated or rural environments | 5 | 3 | “Well, it was good because I knew that if I was [inputting information], and somebody at the other end was seeing that something was really wrong they would have notified the girls in Camrose and would have notified medical assistance immediately. So, it was kind of another area of peace of mind...yeah, we don’t have next door neighbors. We can’t run for help. You know, we’re sort of isolated out here, so like I say, we’re on our own. You kind of just grasp at things, and you watch for signs. But you’re not always fully aware, maybe, of what’s happening until it’s too late”<br>[2/7 participants] | “I don’t know, I didn’t get a full view on what all [the device] could do, simply because it was kind of one-sided. I knew the information was going out, but I didn’t necessarily know what was happening with it. I’m not having that feedback from a medical professional. I feel like it would be a really important piece if this is being used in the future.”<br>[1/7 participants] |
| Improved insight into symptoms | 25 | 7 | “...I felt like I’ve gone crazy for a long time, and it just helps me, even if it’s not my heart rate, and it’s something else. It’s just a peace of mind that it, you know, it’s not my heart.”<br>“I quite like the monitoring aspect from home, just because it gives me a bit more of an insight of what my heart rate is and what it’s doing; just a better overall understanding.”<br>[6/7 participants] | “I think, because I was having so many problems with my heart rate, and then I was tracking it so closely, it did make me hyper-aware of my heart rate a little bit...because now I was really closely monitoring, and it wasn’t going well. So then, I was really concerned about that.<br>[2/7 participants] |
| Concern for privacy | 7 | 7 | “None whatsoever. Like I said, I want to figure out what’s going on, and I don’t want somebody else to have to go through what I’m already dealing with. They might as well use me to figure out what we can do for others, right? So, I have no concerns with it whatsoever.”<br>[6/7 participants] | “It would again come down to what the requirements were for [data sharing]. ... or at the very least had some transparency about what they were doing with it...my information is out and I don’t know what it’s taking off of my phone. What sort of data it’s gathering on me. So that’s something that I see as a limitation of health monitoring.”<br>[1/7 participants] |
| Research potential of devices | 8 | 6 | “Well, I think it was helpful for whoever was tracking to get a better insight into what the aftereffects of COVID does, everybody’s different. It’s a very needed program and when you have somebody who went through what [my spouse] did, it’s really helpful. Maybe it’ll help someone else in [the ESD program]. I think the program is very much needed.”<br>[5/7 participants] | “I’ve heard of studies where because of where the money is coming from, there’s a risk of results in the data being made to fit the needs of the corporation or person or group or whatever. If the data is being used by people who are motivated financially rather than scientifically, and not with the goals of the health of individuals and people.”<br>[1/7 participants] |

### Support in isolated environments

Most felt that wearable devices are likely to improve medical safety for rural communities in the future, possibly through symptom, activity, and vital sign data transmission to clinical teams, family members and caregivers.

### Gaining disease insights

Some patients spontaneously use device data (mostly heart rate) to help them interpret physical symptoms, even without medical guidance. The impacts of biosensor data on mental health were varied. Some found that normal heart rate readings reassured them while others found that elevated heart rates worsened anxiety about their disease.

### Concerns around privacy

Few were concerned data privacy and security. One participant was hesitant to share their data with for-profit device companies, specifically.

### Potential for research

Almost universally, participants felt wearable devices held great promise as tools for clinical research and expressed a willingness to contribute their data to the research community. Participants were motivated by a desire to see advancements in understanding and treatment of post-COVID conditions.

## Discussion

We report excellent feasibility characteristics for wrist-worn fitness trackers in post-COVID disease monitoring in rural areas, including hypothesis generating associations suggesting that biosensor data can deliver disease insights at the individual level. Biosensor data was complex and heterogenous, highlighting the need for machine learning techniques to separate signal from noise. Our qualitative work illustrated a unique lived experience for rural patients with technology and unmet needs for remote monitoring tools that integrate with existing clinical care models to ease transitions from hospital to home.

Compared to existing studies, we observed high levels of protocol adherence. For common disease targets like COPD and CHF, RPM protocol adherence is typically low and decreases over time (Singhal & Cowie, 2020; Stehlik et al., 2020; Wu et al., 2021). Our post-COVID population was younger than most existing RPM cohorts, had relatively few comorbidities and were generally comfortable with technology, perhaps explaining our favorable result. Also, most existing studies RPM protocols use medical-grade devices, which are often unfamiliar to patients and challenging to operate. Consumer choice in technology is key driver of success for RPM and consumer-grade devices that offer non-medical applications present an opportunistic substrate for building scalable RPM platforms (Curry, 2023). The drawback of consumer-grade biosensors is that sensor accuracy is often unknown and cannot be assumed clinical grade (Bent et al., 2020). However, as sensor accuracy and signal detection algorithms improve for longitudinal patient data, we will be able to better compensate for imperfection in underlying discrete data. Learning to process data from consumer-grade biosensors is foundational for just-in-time medical decision making, especially in relatively healthy patient populations who haven’t been prescribed a medical grade device (Takahashi et al., 2022). Our study underscores the need to develop patient-centered technologies built with the non-medical consumer in mind.

Our group level associations and individual case examples highlight the potential for smartwatches to deliver novel disease insights in scientific studies. In our post-COVID patients, anxiety and stress correlated with activity and resting heart rate while physical symptom correlated with sleep. This granular, continuous data paints a rich picture of disease at the individual level, forming a foundation for precision medicine and improving on traditional forms of episodic measurement. Given our small sample size, our observed group-level associations are hypothesis generating only. Subjectively, the heterogeneity seen across individual data supports the hypothesis that post-COVID conditions comprise a range of distinct phenotypes rather than a single entity (Lusczek et al., 2021; Osuchowski et al., 2021), a possibility that will hopefully be further elucidated in ongoing clinical trials that utilize wearable devices (Moore Vogel, 2023). As more studies using wearable biosensors emerge it will be important to create and adhere to standards in metric derivation and validation so that results are clinically applicable.

Our patient interviews provided a rich context for interpreting quantitative data and brought to the fore a range of patient experiences that are key for understanding the future impacts of wearable devices in medicine. Continuous physiologic data can amplify or dampen anxiety around disease depending on the individual. In some instances, normal device data helped to reassure when symptoms were fluctuating unpredictably. For others, abnormal heart rate data worsened anxiety around mild or absent physical sensations. The potential impacts of personalized biodata on mental health are poorly described in existing literature and for some it could contribute to excess healthcare utilization (Rosman et al., 2020). Our result highlights the need to better understand these impacts through further research and to improve medical support patients who choose to wear devices. Interestingly, concerns about data-sharing and privacy were infrequently raised by our patients but are known to be major determinants of success for wearable devices in healthcare, overall (Banerjee et al., 2018; Cilliers, 2020).

Patients unfamiliar with wrist-worn devices in our study required a significant amount of time and support during device set-up. For those who owned a device already, changing to a new one was unpopular, and some patients refused enrollment or withdrew consent as a result. Human factors like this are critical determinants of success for RPM platforms. Shin et al. (2019) found that wearables are best accepted by patients who are motivated to monitor a chronic condition. Yin et al. (2022) reported that convenience, social influence, and expectation of improved health are important for uptake, while cost and perceived risk are less so. In rural settings, such as ours, perceived benefits for wearable technologies might be elevated as in-person services are often inaccessible (reviewed in Brahmbhatt et al., 2022). In future work using smartwatches rather than fitness trackers would likely yield even stronger feasibility data given the attractiveness of their non-medical applications like text messaging, music, and social networking, all of which encourage wear-time. Indeed, many patients in our study were particularly reluctant to use a fitness tracker if they already owned a smartwatch.

### Limitations

Our study has several limitations. Most notably, our sample size was small, and it is unclear if our findings can be replicated a larger population. As conditions changed during our study (vaccines emerged and prevalent viral strains shifted) we observed that the post-COVID phenotype also changed. In the early stages of our work, we observed more severe respiratory symptoms and most patients had been hospitalized, often recovering from a period of mechanical ventilation. In the later stages, patients were often referred from community clinics with more mild respiratory disease but more severe constitutional symptoms (ex. fatigue or poor sleep). The shifting disease phenotype makes it challenging to infer similar results for future endemic states for COVID. Importantly, in this setting our patients were visited near-daily in their homes or via telemedicine by therapists, which could have artificially increased protocol adherence, and critically, a relatively low recruitment rate (60 percent) suggests that population-level impacts for wearable technologies remains uncertain.

### Conclusions

We find promising feasible characteristics for wrist-worn devices in remote disease tracking for post-COVID conditions in rural communities. Our data are foundational for future testing of consumer-grade devices in the medical sphere, highlighting the need for co-created RPM platforms designed with patient/consumer technology preference in mind. Such innovations have great potential to improve healthcare access and safety for otherwise isolated populations.

## Data Availability

All relevant data are within the manuscript and its Supporting Information files.

## Acknowledgements

We acknowledge the extensive efforts of the Camrose Early Supported Discharge team (Dana Norton, Alyssa Rose, Sharene Lamson) in providing clinical and administrative support. We also acknowledge the contributions of the Glenrose research team (Elton Lam, Tod Vandenberg, Stephan Pham) for their provision of analytic support. Funding for this work was generously provided by Covenant Health and Alberta Health Services. No direct funding was received from FitBit.

## Supplementary Materials

**Table S1.** Patient characteristics – complications for hospitalized patients (N=5), comorbid conditions captured with administrative data diagnostic codes (N=10), medications in the year prior to COVID-19 diagnosis generated by WHO ATC coding system, and symptom scores by body system.

| **Complications for Hospitalized Patients** | **Frequency** |
| --- | --- |
| Cardiovascular (ex. acute MI, atrial fibrillation, pulmonary embolism) | 5 |
| Infectious (non-respiratory) (ex. urinary tract infection, sepsis) | 3 |
| Excretory (ex. acute kidney injury) | 2 |
| Metabolic (alkalosis, hypokalemia) | 2 |
| Hematologic (ex. bleeding) | 2 |
| **Medical comorbidities** | **Frequency** |
| Arrhythmia | 1 |
| Pulmonary Circulation Disease | 2 |
| Hypertension | 2 |
| Chronic Pulmonary Disease | 1 |
| Peptic Ulcer Disease | 1 |
| Fluid and Electrolyte Diseases | 1 |
| **Medications** | **Frequency** |
| **Number of Medications** | **4.3 [SD=3.1]** |
| Alimentary Tract | 3 |
| Blood and blood forming | 0 |
| Cardiovascular | 4 |
| Dermatologic | 0 |
| Genitourinary | 3 |
| Hormonal | 5 |
| Anti-Infective | 6 |
| Antineoplastic | 0 |
| Musculoskeletal | 2 |
| Nervous system | 8 |
| Antiparasitic | 0 |
| Respiratory | 5 |
| Sensory | 0 |
| Various (i.e. others) | 0 |
| **Physical Symptoms (Score 0-4, [SD])** |  |
| ***Constitutional (6 symptoms)*** | 1.43[0.83] |
| Fatigue | 2.46 [1.18] |
| Aches and pains | 1.45 [1.32] |
| Muscle weakness | 1.32 [1.02] |
| Poor sleep | 1.72 [0.98] |
| Fever | 0.04 [0.09] |
| Feeling Generally Unwell | 1.60 [1.52] |
| ***Gastrointestinal (3 symptoms)*** | 0.31[0.55] |
| Nausea and Vomiting | 0.34 [0.62] |
| Diarrhea | 0.12 [0.28] |
| Abdominal Pain | 0.47 [0.88] |
| ***Neurological (3 symptoms)*** | 0.61[0.62] |
| Headache | 1.50 [1.37] |
| Loss of taste | 0.17 [0.47] |
| Loss of smell | 0.16 [0.51] |
| ***Respiratory (4 symptoms)*** | 0.90[0.55] |
| Cough | 0.92 [0.96] |
| Shortness of Breath | 1.70 [1.12] |
| Runny Nose | 0.54 [0.54] |
| Sore Throat | 0.47 [0.69] |
| ***Cardiovascular (2 symptoms)*** | 0.68[0.90] |
| Chest Pain | 0.83 [1.12] |
| Palpitations | 0.54 [0.75] |
| **Mental health Symptoms** |  |
| General Anxiety Disorder-7 (out of 21) | 16.3 [7.0] |
| Patient Health Questionnaire-9 (out of 27) | 20.2 [8.2] |
| Perceived Stress Scale (out of 40) | 29.7 [5.3] |
| **Baseline technology and health literacy scores** |  |
| Tech comfort (7-items, avg score [0-5]) | 3.59 |
| Health Literacy (4-items, avg score [0-5]) | 3.68 |

**Table S2.** Mean difference in average symptoms scores for weeks 1-4 vs. 8+ for patients participating for more than 8 weeks (N=5).

| Symptom | Mean difference after 8 weeks | P-value |
| --- | --- | --- |
| Constitutional | 0.18 | 0.63 |
| Neuro | 0.20 | 0.37 |
| Gastrointestinal | -0.03 | 0.17 |
| Respiratory | -0.23 | 0.03 |
| Cardiovascular | -0.39 | 0.09 |
| Mood | -2.81 | 0.66 |
| Anxiety | -2.96 | 0.86 |
| Stress | -4.36 | 0.45 |
| Total Steps | -108.4 | 0.18 |
| Resting Heart Rate (bpm) | -4.31 | 0.32 |
| Average Heart Rate (bpm) | -5.99 | 0.10 |

**Table S3:** Correlation coefficients between device wear-time (pooled weekly) and patient characteristics, symptom severity, technology acceptance and time under observation (N=10).

| Factor | Correlation | P-value |
| --- | --- | --- |
| Age | -0.27 | 0.46 |
| Admission Length | -0.38 | 0.28 |
| Symptoms Survey Response Rate | 0.67 | 0.03 |
| Mental Health Survey Response Rate | 0.53 | 0.11 |
| Tech Readiness Scores | 0.38 | 0.28 |
| All Symptoms | -0.43 | 0.21 |
| Generalized Anxiety Disorder-7 | -0.59 | 0.07 |
| Patient Health Questionnaire 9 | -0.52 | 0.13 |
| Perceived Stress Scale | -0.61 | 0.06 |

**Figure S1.**
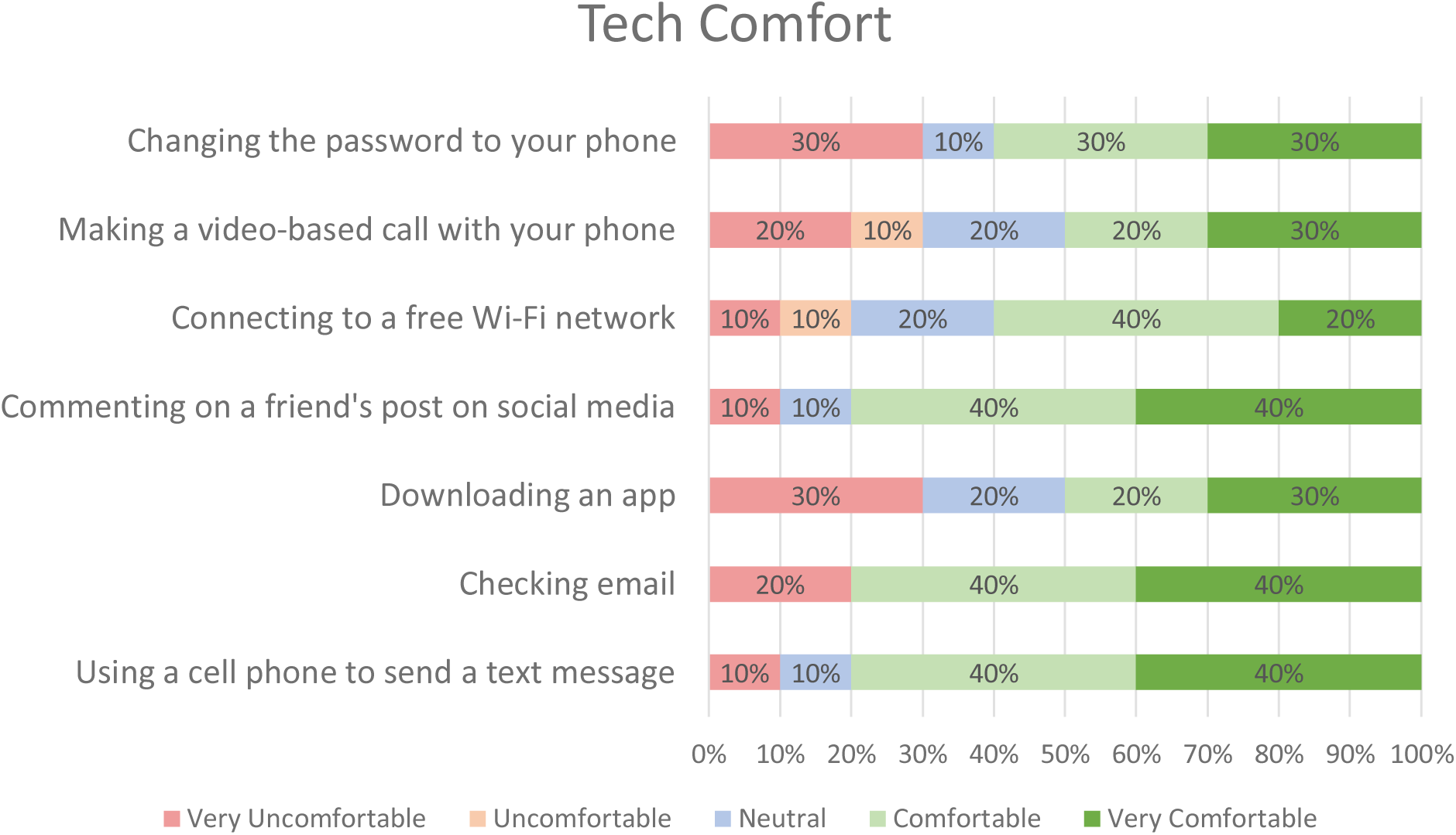
Baseline technology comfort survey responses (N=10).

**Figure S2.**
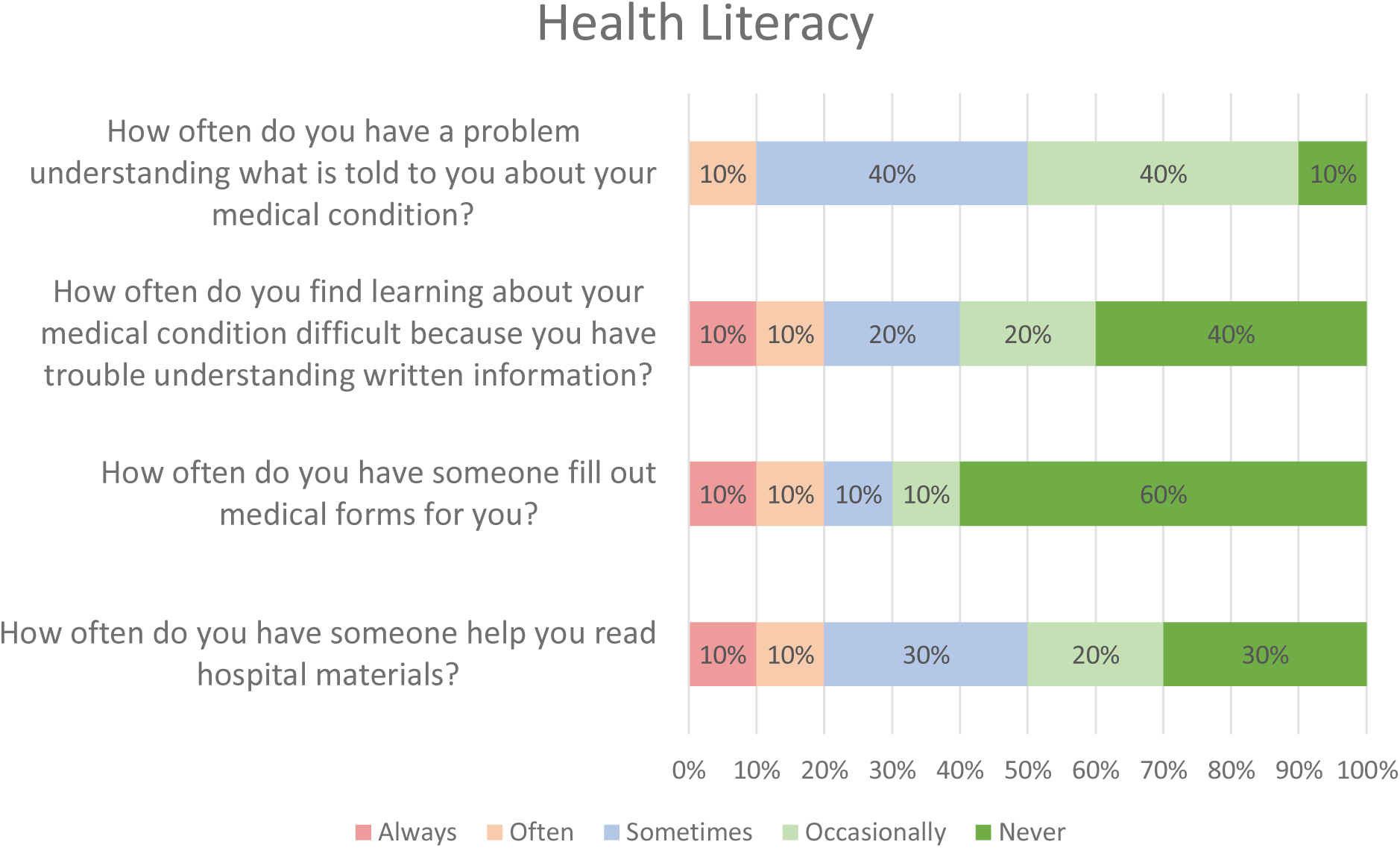
Baseline health literacy survey responses (N=10)

## References

1. Banerjee, S. (Sy), Hemphill, T., & Longstreet, P. (2018). Wearable devices and healthcare: Data sharing and privacy. The Information Society, 34(1), 49–57. 10.1080/01972243.2017.1391912

2. Bent, B., Goldstein, B. A., Kibbe, W. A., & Dunn, J. P. (2020). Investigating sources of inaccuracy in wearable optical heart rate sensors. Npj Digital Medicine, 3(1), 18. 10.1038/s41746-020-0226-6

3. Brahmbhatt, D. H., Ross, H. J., & Moayedi, Y. (2022). Digital Technology Application for Improved Responses to Health Care Challenges: Lessons Learned From COVID-19. The Canadian Journal of Cardiology, 38(2), 279–291. 10.1016/j.cjca.2021.11.014

4. Canali, S., Schiaffonati, V., & Aliverti, A. (2022). Challenges and recommendations for wearable devices in digital health: Data quality, interoperability, health equity, fairness. PLOS Digital Health, 1(10), e0000104. 10.1371/journal.pdig.0000104

5. Channa, A., Popescu, N., Skibinska, J., & Burget, R. (2021). The Rise of Wearable Devices during the COVID-19 Pandemic: A Systematic Review. Sensors, 21(17), 5787. 10.3390/s21175787

6. Chouliara, N., Cameron, T., Byrne, A., Lewis, S., Langhorne, P., Robinson, T., Waring, J., Walker, M., & Fisher, R. (2023). How do stroke early supported discharge services achieve intensive and responsive service provision? Findings from a realist evaluation study (WISE). BMC Health Services Research, 23(1), 299. 10.1186/s12913-023-09290-1

7. Cilliers, L. (2020). Wearable devices in healthcare: Privacy and information security issues. Health Information Management Journal, 49(2–3), 150–156. 10.1177/1833358319851684

8. Cohen, S., Kamarck, T., & Mermelstein, R. (1983). A Global Measure of Perceived Stress. Journal of Health and Social Behavior, 24(4), 385. 10.2307/2136404

9. Davis, H. E., McCorkell, L., Vogel, J. M., & Topol, E. J. (2023). Long COVID: Major findings, mechanisms and recommendations. Nature Reviews Microbiology, 21(3), 133–146. 10.1038/s41579-022-00846-2

10. Fraser, M. J., Gorely, T., O’Malley, C., Muggeridge, D. J., Giggins, O. M., & Crabtree, D. R. (2022). Does Connected Health Technology Improve Health-Related Outcomes in Rural Cardiac Populations? Systematic Review Narrative Synthesis. International Journal of Environmental Research and Public Health, 19(4), 2302. 10.3390/ijerph19042302

11. Harris, P. A., Taylor, R., Thielke, R., Payne, J., Gonzalez, N., & Conde, J. G. (2009). Research electronic data capture (REDCap)—a metadata-driven methodology and workflow process for providing translational research informatics support. Journal of biomedical informatics, 42(2), 377–381.

12. Iqbal, S. M. A., Mahgoub, I., Du, E., Leavitt, M. A., & Asghar, W. (2021). Advances in healthcare wearable devices. Npj Flexible Electronics, 5(1), 9. 10.1038/s41528-021-00107-x

13. Kang, H. S., & Exworthy, M. (2022). Wearing the Future-Wearables to Empower Users to Take Greater Responsibility for Their Health and Care: Scoping Review. JMIR mHealth and uHealth, 10(7), e35684. 10.2196/35684

14. Khondakar, K. R., & Kaushik, A. (2022). Role of Wearable Sensing Technology to Manage Long COVID. Biosensors, 13(1), 62. 10.3390/bios13010062

15. Koc, H. C., Xiao, J., Liu, W., Li, Y., & Chen, G. (2022). Long COVID and its Management. International Journal of Biological Sciences, 18(12), 4768–4780. 10.7150/ijbs.75056

16. Kroenke, K., Spitzer, R. L., & Williams, J. B. W. (2001). The PHQ-9: Validity of a brief depression severity measure. Journal of General Internal Medicine, 16(9), 606–613. 10.1046/j.1525-1497.2001.016009606.x

17. Kwok, E. S. H., Clapham, G., & Calder-Sprackman, S. (2021). The Impact of COVID-19 Pandemic on Emergency Department Visits at a Canadian Academic Tertiary Care Center. The Western Journal of Emergency Medicine, 22(4), 851–859. 10.5811/westjem.2021.2.49626

18. Liao, Y., Thompson, C., Peterson, S., Mandrola, J., & Beg, M. S. (2019). The Future of Wearable Technologies and Remote Monitoring in Health Care. American Society of Clinical Oncology Educational Book. American Society of Clinical Oncology. Annual Meeting, 39, 115–121. 10.1200/EDBK_238919

19. Lu, L., Zhang, J., Xie, Y., Gao, F., Xu, S., Wu, X., & Ye, Z. (2020). Wearable Health Devices in Health Care: Narrative Systematic Review. JMIR mHealth and uHealth, 8(11), e18907. 10.2196/18907

20. Lusczek, E. R., Ingraham, N. E., Karam, B. S., Proper, J., Siegel, L., Helgeson, E. S., Lotfi-Emran, S., Zolfaghari, E. J., Jones, E., Usher, M. G., Chipman, J. G., Dudley, R. A., Benson, B., Melton, G. B., Charles, A., Lupei, M. I., & Tignanelli, C. J. (2021). Characterizing COVID-19 clinical phenotypes and associated comorbidities and complication profiles. PLOS ONE, 16(3), e0248956. 10.1371/journal.pone.0248956

21. Mishra, T., Wang, M., Metwally, A. A., Bogu, G. K., Brooks, A. W., Bahmani, A., Alavi, A., Celli, A., Higgs, E., Dagan-Rosenfeld, O., Fay, B., Kirkpatrick, S., Kellogg, R., Gibson, M., Wang, T., Hunting, E. M., Mamic, P., Ganz, A. B., Rolnik, B., … Snyder, M. P. (2020). Pre-symptomatic detection of COVID-19 from smartwatch data. Nature Biomedical Engineering, 4(12), 1208–1220. 10.1038/s41551-020-00640-6

22. Osuchowski, M. F., Winkler, M. S., Skirecki, T., Cajander, S., Shankar-Hari, M., Lachmann, G., Monneret, G., Venet, F., Bauer, M., Brunkhorst, F. M., Weis, S., Garcia-Salido, A., Kox, M., Cavaillon, J.-M., Uhle, F., Weigand, M. A., Flohé, S. B., Wiersinga, W. J., Almansa, R., … Rubio, I. (2021). The COVID-19 puzzle: Deciphering pathophysiology and phenotypes of a new disease entity. The Lancet. Respiratory Medicine, 9(6), 622–642. 10.1016/S2213-2600(21)00218-6

23. Parmanto, B., Lewis, A. N., Graham, K. M., & Bertolet, M. H. (2016). Development of the Telehealth Usability Questionnaire (TUQ). International Journal of Telerehabilitation, 8(1), 3–10. 10.5195/ijt.2016.6196

24. Rodriguez-León, C., Villalonga, C., Munoz-Torres, M., Ruiz, J. R., & Banos, O. (2021). Mobile and Wearable Technology for the Monitoring of Diabetes-Related Parameters: Systematic Review. JMIR mHealth and uHealth, 9(6), e25138. 10.2196/25138

25. Rosman, L., Gehi, A., & Lampert, R. (2020). When smartwatches contribute to health anxiety in patients with atrial fibrillation. Cardiovascular Digital Health Journal, 1(1), 9–10. 10.1016/j.cvdhj.2020.06.004

26. Shin, G., Jarrahi, M. H., Fei, Y., Karami, A., Gafinowitz, N., Byun, A., & Lu, X. (2019). Wearable activity trackers, accuracy, adoption, acceptance and health impact: A systematic literature review. Journal of Biomedical Informatics, 93, 103153. 10.1016/j.jbi.2019.103153

27. Singhal, A., & Cowie, M. R. (2020). The Role of Wearables in Heart Failure. Current Heart Failure Reports, 17(4), 125–132. 10.1007/s11897-020-00467-x

28. Smuck, M., Odonkor, C. A., Wilt, J. K., Schmidt, N., & Swiernik, M. A. (2021). The emerging clinical role of wearables: Factors for successful implementation in healthcare. Npj Digital Medicine, 4(1), 45. 10.1038/s41746-021-00418-3

29. Spitzer, R. L., Kroenke, K., Williams, J. B. W., & Löwe, B. (2006). A Brief Measure for Assessing Generalized Anxiety Disorder: The GAD-7. Archives of Internal Medicine, 166(10), 1092. 10.1001/archinte.166.10.1092

30. Stehlik, J., Schmalfuss, C., Bozkurt, B., Nativi-Nicolau, J., Wohlfahrt, P., Wegerich, S., Rose, K., Ray, R., Schofield, R., Deswal, A., Sekaric, J., Anand, S., Richards, D., Hanson, H., Pipke, M., & Pham, M. (2020). Continuous Wearable Monitoring Analytics Predict Heart Failure Hospitalization: The LINK-HF Multicenter Study. Circulation: Heart Failure, 13(3), e006513. 10.1161/CIRCHEARTFAILURE.119.006513

31. Takahashi, S., Nakazawa, E., Ichinohe, S., Akabayashi, A., & Akabayashi, A. (2022). Wearable Technology for Monitoring Respiratory Rate and SpO2 of COVID-19 Patients: A Systematic Review. Diagnostics, 12(10), 2563. 10.3390/diagnostics12102563

32. Wu, C.-T., Li, G.-H., Huang, C.-T., Cheng, Y.-C., Chen, C.-H., Chien, J.-Y., Kuo, P.-H., Kuo, L.-C., & Lai, F. (2021). Acute Exacerbation of a Chronic Obstructive Pulmonary Disease Prediction System Using Wearable Device Data, Machine Learning, and Deep Learning: Development and Cohort Study. JMIR mHealth and uHealth, 9(5), e22591. 10.2196/22591

33. Yin, Z., Yan, J., Fang, S., Wang, D., & Han, D. (2022). User acceptance of wearable intelligent medical devices through a modified unified theory of acceptance and use of technology. Annals of Translational Medicine, 10(11), 629. 10.21037/atm-21-5510

